# Perceptual super-resolution in multiple sclerosis MRI

**DOI:** 10.1101/2024.08.02.24311394

**Authors:** Diana L. Giraldo, Hamza Khan, Gustavo Pineda, Zhihua Liang, Alfonso Lozano, Bart Van Wijmeersch, Henry C. Woodruff, Philippe Lambin, Eduardo Romero, Liesbet M. Peeters, Jan Sijbers

**Affiliations:** imec-Vision Lab, University of Antwerp, Antwerp, Belgium.; Computer Imaging and Medical Applications Laboratory - CimLab, Universidad Nacional de Colombia, Bogotá, Colombia; University MS Center, Biomedical Research Institute, Hasselt University, Belgium; Data Science Institute (DSI), Hasselt University, Belgium; The D-Lab, Department of Precision Medicine, GROW-Research Institute for Oncology and Reproduction, Maastricht University, Netherlands; Department of Diagnostic Imaging, Hospital Universitario Nacional, Universidad Nacional de Colombia, Bogota, Colombia; Noorderhart, Revalidatie en MS, Pelt, Belgium; Department of Radiology and Nuclear Imaging, GROW-Research Institute for Oncology and Reproduction, Maastricht University Medical Centre+, Maastricht, Netherlands.; µNEURO Research Center of Excellence, University of Antwerp, Antwerp, Belgium

**Keywords:** Super Resolution, MRI, Multiple Sclerosis, Lesion Segmentation, CNN, fine-tuning

## Abstract

Magnetic resonance imaging (MRI) is crucial for diagnosing and monitoring of multiple sclerosis (MS) as it is used to assess lesions in the brain and spinal cord. However, in real-world clinical settings, MRI scans are often acquired with thick slices, limiting their utility for automated quantitative analyses. This work presents a single-image super-resolution (SR) reconstruction framework that leverages SR convolutional neural networks (CNN) to enhance the through-plane resolution of structural MRI in people with MS (PwMS). Our strategy involves the supervised fine-tuning of CNN architectures, guided by a content loss function that promotes perceptual quality, as well as reconstruction accuracy, to recover high-level image features. Extensive evaluation with MRI data of PwMS shows that our SR strategy leads to more accurate MRI reconstructions than competing methods. Furthermore, it improves lesion segmentation on low-resolution MRI, approaching the performance achievable with high-resolution images. Results demonstrate the potential of our SR framework to facilitate the use of low-resolution retrospective MRI from real-world clinical settings to investigate quantitative image-based biomarkers of MS.

## 1 Introduction

Multiple sclerosis (MS) is a chronic autoimmune neurodegenerative disease characterized by inflammation and demyelination of nerve axons in the central nervous system. This damage leads to the formation of lesions, which are the most important markers of disease activity [1]. Diagnosis and monitoring of people with MS (PwMS) relies on the acquisition of Magnetic Resonance Imaging (MRI), particularly T2-weighted (T2-W) fluid-attenuated inversion recovery (FLAIR), to assess lesions in the white matter (WM) [2]. Although current clinical guidelines recommend acquiring high-resolution (HR) T2-W FLAIR MRI using 3D sequences [2], in clinical settings images have been often acquired with 2D sequences, where the resulting ’3D’ image is rather a stack of thick 2D slices with highly anisotropic voxels. Such multi-slice images are faster to acquire, are less prone to motion artifacts, and their in-plane resolution is often sufficient for visual inspection by radiologists. However, their poor through-plane resolution hampers their use for precise quantitative analyses (e.g., radiomics) as most of the automated methods proposed for lesion segmentation and morphometric analyses require HR images with isotropic voxels [3, 4, 5]. In this scenario, super-resolution (SR) methods aiming to improve the spatial resolution of acquired MRI would facilitate the use of real-world MRI and clinical data of PwMS to investigate MS biomarkers.

Super-resolution methods aim to estimate an unknown HR MRI from one or more acquired low-resolution (LR) MRIs. Model-based methods assume an explicit imaging model of the MRI acquisition and seek a numerical solution of the ill-posed inverse problem by introducing regularization terms to constrain the solution space [6, 7, 8, 9]. These model-based SR methods, however, require multiple multi-slice LR images to reconstruct one HR MRI. Hence, they are not suitable for single-image SR. Even when two multi-slice LR images (e.g. with orthogonal slice direction) are available, their performance is limited [10]. Alternatively, learning-based single-image SR approaches, for example based on convolutional neural networks (CNN), have demonstrated impressive performance in natural images, due to their ability to learn the relation between the LR and HR images from data [11, 12, 13, 14, 15, 16]. A straightforward procedure is to train a CNN generative model with paired LR-HR samples, enabling the model to learn a SR mapping that can be applied to unseen LR data with generally fast inference times. Despite their demonstrated effectiveness on natural images, the performance of these SR approaches heavily relies on the variability and characteristics of the data the models were trained and tested with. CNN models trained with large datasets of natural images do not capture the imaging conditions, artifacts, anatomical structures, and pathological features present in medical images. Therefore their application to medical images within certain contexts requires the models to be trained or fine-tuned with data containing such domain-specific features.

In the context of brain MRI SR, several works have presented and evaluated CNNs for single-image SR of structural MRI sequences, primarily T1-W and T2-W scans. Sanchez and Vilaplana [17] proposed a 3D Generative Adversarial Network (GAN) architecture inspired by the SRGAN model [13]. Their method achieved promising quantitative results and demonstrated the potential of GANs for MRI SR compared to conventional cubic spline interpolation. Pham *et al.* [18] presented a 3D CNN based on residual learning, with the underlying assumption that it is easier to find a mapping from the missing high-frequency information to HR, instead of finding a direct mapping from LR to HR. A similar residual learning approach was adopted by Du *et al.* [19], who proposed a 2D CNN network for SR reconstruction of multi-slice T1-W and T2-W MRI, although it was mainly evaluated on synthetic brain images. Addressing the challenge of collecting training data, Zhao *et al.* [20, 21] introduced a self-supervised approach based on 2D CNNs, which leveraged the high in-plane resolution of acquired MRI to train a SR CNN for increasing the through-plane resolution. This method showcased improved performance over previous self-supervised methods when evaluated for the SR of multi-slice T2-W MRI. However, the requirement to train or finetune a CNN model each time the method is applied to a new image poses an important practical limitation in terms of computational cost and processing time. A CNN model trained to convert a brain MRI of any orientation, resolution and contrast into an HR T1-W MRI was recently presented [22, 23]. While this approach holds promise for facilitating standard morphometric analyses by inpainting normal-appearing tissue in pathological areas, it does not facilitate the quantitative analysis of lesions in PwMS.

Most CNN strategies for MRI SR rely on minimizing either the mean squared error (*L*_2_ loss) [24, 18, 19] or mean absolute error (*L*_1_ loss) [21, 23] between the model output and the ground truth HR image during model training. While this minimization approach results in improved reconstruction accuracy measures such as peak signal-to-noise ratio (PSNR) and structural similarity (SSIM), it can produce images with over-smoothed textures and blurry boundaries [25, 15]. To address this limitation, previous works have explored the use of GANs [17, 26], which encourage the model to generate more realistic-looking images by incorporating an adversarial loss. However, this adversarial approach carries the risk of hallucinating structures or introducing artifacts in generated images. Another promising approach to improve the perceptual quality in SR is the use of loss functions that compare high-level image features rather than relying solely on pixel-wise similarities. These high-level features are often extracted from intermediate layers of pre-trained neural networks [12]. Perceptual losses, which transfer semantic knowledge from the pre-trained loss network, have demonstrated their effectiveness in improving the perceptual quality of single-image SR for natural images [12, 27]. Recently, Zhang *et al.* [26] incorporated a perceptual loss into a GAN framework for MRI SR, which was trained with T1-W brain MRI of PwMS and then tested on T2-W FLAIR.

Current state-of-the-art approaches for single-image SR of multi-slice MRI [19, 21, 26] have been evaluated in scenarios where the upsampling scale factor between the slice thickness of LR and HR ranges from 2 to 6. As expected, the performance of SR reconstruction decreases as the input slice thickness increases, leading to greater challenges in faithfully recovering anatomical structures and details. The larger resolution gap between LR input and HR target exacerbates the risk of hallucinating artificial features, compromising the diagnostic quality and reliability of SR outputs, a critical concern when applying SR to brain MRI of PwMS. A particularly challenging scenario, that is prevalent in real-world retrospective image datasets of PwMS, involves MRI acquisitions with a thick slice spacing of 6 mm. Moreover, these multi-slice MRI scans are often acquired with slice gaps, a factor that is frequently overlooked when generating LR training data for SR models but can significantly influence their performance [28].

In this paper, we present a SR framework that leverages SR CNN architectures to enhance the resolution of multi-slice structural MRI of PwMS, reducing the slice spacing from 6 mm to 1mm. Our strategy involves fine-tuning SR models with image patches extracted from T2-W FLAIR and T1-W MRI data of PwMS. The fine-tuning of SR models is guided by a content loss *L*_c_ which simultaneously promotes perceptual quality and pixel-wise reconstruction accuracy, ensuring realistic textures and well-defined tissue boundaries in the reconstructed HR images. Our framework reconstructs HR MRI volumes from single LR inputs, enabling more accurate downstream 3D analysis. Through a comprehensive evaluation using multi-center MRI data, we demonstrate that our SR framework outperforms existing methods in terms of reconstruction accuracy, and additionally improves the performance of automated lesion segmentation on T2-W FLAIR MRI, a highly relevant task in the context of multiple sclerosis.

We summarize the contributions of this work as follow:

- We present a single-image SR framework for multi-slice MRI based on the adaptation of CNN architectures to the domain of structural MRI in MS.
- We fine-tune two state-of-the-art SR CNN models, namely the EDSR [14] and the RealESRGAN [16], using a perceptual loss to recover realistic features, and the mean absolute error (*L*_1_ loss) to control the reconstruction accuracy.
- We incorporate information from the MRI physics model by simulating LR data from HR MRI with an acquisition model that accounts for a slice selection profile, including slice gap, commonly found in clinical multi-slice MRI data.
- We evaluate our framework for MRI SR reconstruction and compare its performance against existing MRI SR methods using MRI datasets of PwMS from different centers.
- We evaluate the impact of our SR strategy in a relevant downstream task: the automated segmentation of white matter lesions on reconstructed T2W-FLAIR images.

We named our framework PRETTIER, a name encapsulating its purpose: ”Perceptual superREsoluTion in mulTIple sclERosis”. The code to apply PRETTIER is available at https://github.com/diagiraldo/PRETTIER.

## 2 Materials and Methods

An overview of the workflow is presented in Figure 1. The process begins with MRI data preparation, consisting of LR MRI simulation and extraction of paired LR-HR patches. Then, in the learning step, CNN models for image SR are fine-tuned using these patches, with final weights selected based on the minimum content loss in a validation set. We then use each fine-tuned CNN model to reconstruct HR MRI volumes from single LR MRI inputs, combining outputs from applying the model in different slice directions. The evaluation step uses an independent set of structural MRI from PwMS and comprises three parts: First, we evaluate the fine-tuning of CNN models with MRI patches. Second, we evaluate the SR framework by reconstructing MRI volumes. Finally, we assess the impact of SR reconstruction on the automated segmentation of WM lesions.

**Figure 1:**
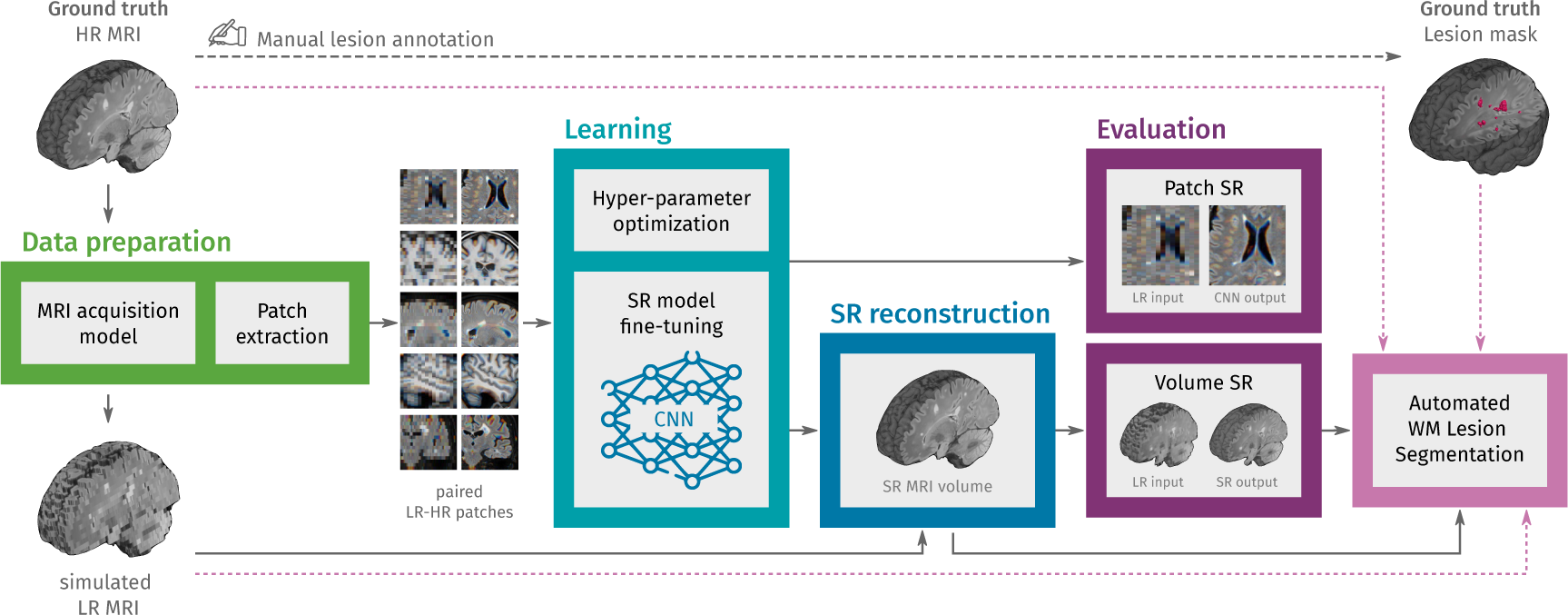
Methodology overview. In the first step, LR multi-slice MRI acquisitions are simulated using ground truth HR MRIs, and paired LR-HR image patches are extracted. In the learning step, pre-trained convolutional neural network (CNN) models for natural image super-resolution (SR) are fine-tuned with the extracted MRI patches. Then, the fine-tuned models are used to reconstruct HR MRI volumes from single LR MRI inputs. In the evaluation step, the SR performance is assessed at the patch level and the MRI volume level. Finally, we also evaluate the effect of SR on the automated segmentation of white matter (WM) lesions.

### 2.1 MRI data

We gathered six different datasets containing 429 HR structural MRIs, T2-W FLAIR and T1-W, of 192 PwMS. The numbers of unique subjects, images and data partition are shown in Table 1.

- **ISBI2015** [29]: 61 T1-W images from the test data for the longitudinal MS lesion segmentation challenge held during the ISBI 2015 conference. All images were acquired on a 3T MRI scanner (Philips).
- **Lesjak-3D** [30]: 30 T2-W FLAIR images accompanied by their lesion annotation resulting from the consensus of three experts. Images were acquired on a 3T Siemens Magnetom Trio MR system at the University Medical Center Ljubljana.
- **MSSEG1** [31]: 53 T2-W FLAIR and 53 T1-W images from the MS lesions segmentation challenge held in MICCAI 2016. For a subset of 15 subjects (subset A) the dataset also provides the WM lesion annotation resulting from the consensus of seven experts. Images were acquired in four different centers with 1.5T and 3T MR scanners from different vendors (Siemens, Philips, and GE).
- **MSSEG2** [32]: 80 T2-W FLAIR images from the MS new lesions segmentation challenge held in MICCAI 2021. This dataset contains scans of 40 PwMS acquired in two different time-points. Images were acquired with 11 different 1.5T and 3T MR scanners.
- **MSPELT**: 56 T2-W FLAIR and 41 T1-W images of 41 PwMS from the Noorderhart Revalidatie & MS Centrum in Pelt, Belgium. All images were acquired in a 1.5T MR scanner (Philips Achieva dStream). The use of this pseudonymized retrospective dataset was approved by the ethical commission of the University of Hasselt.
- **HUN**: 28 T2-W FLAIR and 27 T1-W images of 14 PwMS from the Hospital Universitario Nacional in Bogota, Colombia. This dataset contains, for each subject, scans at two different visits and the manual annotation of WM lesions verified by an expert neuroradiologist (20 years of experience) for each visit. All images were acquired in a 1.5T MR scanner (Philips Multiva) and collected given free consent and ethical approval from the local ethics committee.

**Table 1:** Overview of the multi-centric dataset used in this work. The two datasets used for evaluation, MSSEG1 A and HUN, have manual delineation of lesions to evaluate automated lesion segmentation.

| Dataset | Source | # subjects | # FLAIR images | # T1 images | Lesion annotation | Data partition |
| --- | --- | --- | --- | --- | --- | --- |
| ISBI 2015 | public | 14 | - | 61 | - | Learning |
| Lesjak-3D | public | 30 | 30 | - | ✓ | Learning |
| MSSEG1 B | public | 38 | 38 | 38 | - | Learning |
| MSSEG2 | public | 40 | 80 | - | - | Learning |
| MSPELT | clinical | 41 | 56 | 41 | - | Learning |
| MSSEG1 A | public | 15 | 15 | 15 | ✓ | Evaluation |
| HUN | clinical | 14 | 28 | 27 | ✓ | Evaluation |
| Total |  | 192 | 247 | 182 |  |  |

Detailed information about data acquisition and compliance with ethical standards for the four public datasets (ISBI2015, Lesjak-3D, MSSEG-1, and MSSEG-2) can be found in their corresponding publications.

#### 2.1.1 MRI pre-processing

The raw versions of HR T2-w FLAIR and T1-W images were denoised with adaptive non-local means [33], and bias-field corrected with N4 algorithm [34]. For each image, a brain mask was estimated using the HD-BET tool [35]. All images, brain masks and ground truth lesion masks were adjusted to have isotropic voxels of 1 mm^3^ using cubic interpolation for MRIs and nearest neighbor interpolation for the binary masks.

#### 2.1.2 Data partition

The set of 429 MRIs was split into learning (204 T2-W FLAIR and 140 T1-W MRIs) and evaluation (43 T2-W FLAIR and 42 T1-W MRIs) as shown in Table 1. Images in the learning set were further partitioned into training and validation with a proportion of 7:3. Images in the validation set were used to optimize training hyper-parameters and select the best model weights during fine-tuning.

### 2.2 Data preparation

#### 2.2.1 Simulation of LR images

We obtained pairs of LR-HR MRI volumes (**y_i_**, **x**) by applying the multi-slice MRI acquisition model *M_i_*[7] to the HR ground truth image:

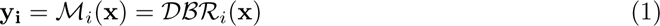

Where *R_i_* is a rotation operator that accounts for the slice direction of the multi-slice LR image, *B* is the blurring operator with a filter that accounts for the slice-selection profile, and *D* is the downsampling operator. For each HR image **x**, we simulated three LR images *{y_i_}*^3^ with orthogonal slice orientations (axial, sagittal, and coronal), with slice thickness of 5 mm and slice spacing of 6 mm (a slice gap of 1 mm). The simulated LR images have highly anisotropic voxels of 1*×*1*×*6 mm^3^, which are common in retrospective clinical multi-slice MRI.

#### 2.2.2 Extraction of patches

Most CNN models for image SR receive as inputs 2D images with 3 color channels (R: red, G: green, B: blue). For model fine-tuning, we randomly extracted pairs of 3-channel patches from each pair of LR-HR MRI by taking triplets of 2D patches that are contiguous in the third dimension. As we considered only fully convolutional CNN models, there are no restrictions in terms of patch size. To keep the memory requirements relatively low, we extracted HR patches of 96*×*96 while ensuring most of the patch area correspond to brain tissue. Therefore, the LR patches where 96*×*16 or 16*×*96, depending on the slicing orientation they were taken from their corresponding LR volume.

For the learning stage, 18 pairs of LR-HR patches were extracted from each LR-HR MRI pair resulting in 13344 training samples and 3168 validation samples. For the evaluation stage, 9 patch pairs extracted from each MRI pair, resulting in 2040 pairs of LR-HR patches.

### 2.3 Learning

The learning stage consists of fine-tuning two CNN models that have shown excellent performance for SR of natural images. The first model is the *Enhanced Deep Residual Networks for Single Image Super-Resolution* (EDSR) [14], which ranked first in the NTIRE 2017 super-resolution challenge [25]. The second model is the *Real Enhanced Super-Resolution Generative Adversarial Network* (RealESRGAN) [16], which is an improved version of the ESRGAN [27]. The ESRGAN achieved the best perceptual index in the 2018 PIRM challenge on perceptual image super-resolution [15].

#### 2.3.1 CNN architectures

- **EDSR** [14]: it builds upon the SRResNet [13] by removing the batch normalization from residual blocks and adding residual scaling to stabilize training. These modifications allow the use of more filters to improve performance without increasing the required computational resources. We used the architectural configuration and pre-trained weights provided by the authors^1^.
- **RealESRGAN** [16]: its generator is composed by residual-in-residual dense blocks, similar as in ESRGAN [27], but the discriminator is a U-Net that provides per-pixel feedback [36]. We used the architectural configuration and pre-trained weights provided by the authors^2^.

#### 2.3.2 Loss function

The content loss *L*_c_ or objective function to minimize during fine-tuning of the CNN models is the sum of the perceptual loss *L*_perceptual_ and the *L*_1_ loss, therefore:

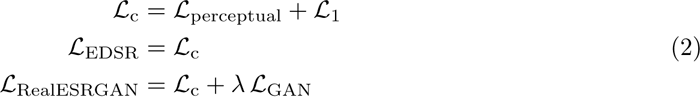

The perceptual loss *L*_perceptual_ compares high-level visual features between the model output and the ground truth, rather than pixel-wise differences. These features can be extracted from a pre-trained network, allowing the knowledge transfer from this loss network to the SR CNN [12]. Here, we calculate the perceptual loss using five layers of a pre-trained VGG-19 [37], following the approach used in the initial training of the RealESRGAN [16]. The adversarial loss *L*_GAN_, used during RealESRGAN fine-tuning, is given by the binary cross entropy with sigmoid function.

#### 2.3.3 Other training details

Both models, EDSR and RealESRGAN, were fine-tuned using the ADAM optimizer [38] and cosine annealing with warm restarts [39] as learning rate scheduler to prevent over-fitting. The batch size was set to 6. Optimal training hyperparameters were selected using the tree-structured parzen estimator [40] within the Optuna [41] framework with the content loss *L*_c_ in the validation set as objective function. The initial learning rate for EDSR was 2*×*10*^−^*^4^ with scheduler parameters *T*_0_ = 8 and *T*_mult_ = 2. In the case of RealESRGAN, initial learning rates and scheduler parameters were tuned for the generator and discriminator. The generator initial learning rate was 1*×*10*^−^*^4^ with scheduler parameters *T*_0_ = 6 and *T*_mult_ = 1, the discriminator initial learning rate was 5*×*10*^−^*^6^ with scheduler parameters *T*_0_ = 8 and *T*_mult_ = 2. The adversarial loss weight *λ* for RealESRGAN fine-tuning was 0.05. Fine-tuning was run for 100 epochs, and the best weights for each model were selected based on the minimum content loss *L*_c_ in the validation set.

### 2.4 SR reconstruction of MRI volumes

After fine-tuning a CNN model for SR of MRI patches, the next step is to leverage the model for reconstructing HR MRI volumes from single LR MRI inputs. For this purpose, we applied the fine-tuned model to LR slices along each of the two in-plane dimensions using a sliding window approach. Specifically, we take 3 contiguous LR slices as the 3-channel input of the SR model, then we combine the HR outputs corresponding to the same slice location using a weighted average. This sliding window approach mitigates the stacking artifacts that could arise from single-slice inference. As this process is done for each in-plane dimension, the result is a pair of volumes consisting of stacked HR slices. These intermediate volumes are then averaged to produce the final SR reconstructed MRI volume.

### 2.5 Evaluation

We evaluated MRI SR at two levels: first, a patch-based assessment to evaluate the fine-tuned SR CNN models on unseen MRI patches. Second, a volume-based assessment to evaluate the SR reconstruction of MRI volumes. Additionally, we apply automated lesion segmentation methods to SR reconstructed T2-W FLAIR MRIs to assess the impact of our SR approach on downstream tasks.

#### 2.5.1 SR of MRI patches

For the patch-based assessment, we used paired LR-HR patches extracted from MRIs in the evaluation set. We compared the output of fine-tuned SR models applied to LR patches against ground truth HR patches using an extended set of image quality measures. These included the widely used PSNR and SSIM, as well as five additional measures:

- Visual information fidelity (VIF) [42], which combines the reference image information and the mutual information between the reference and the distorted image. Its calculation relies on a statistical model for natural scenes, a model for image distortions, and a model of the human visual system.
- Feature similarity index (FSIM) [43], which characterizes the image local quality by combining the image phase congruency and the image gradient magnitude.
- Visual saliency-induced index (VSI) [44], which uses a visual saliency map as a feature to characterize local quality and as a weighting factor when combining it with gradient and chrominance feature maps.
- Haar perceptual similarity index (HaarPSI) [45], which utilizes both high- and low-frequency Haar wavelet coefficients to assess local similarities and weigh local importance. It can be seen as a simplification of the FSIM.
- Deep image structure and texture similarity (DISTS) [46], which combines texture similarity and structure similarity, both computed with feature maps extracted from a pre-trained VGG16.

While PSNR and SSIM compare pixel-wise accuracy, the five additional metrics evaluate visual features extracted from the images using hand-crafted filters or pre-trained CNNs. Recent studies [47, 48] show that these visual feature metrics (VIF, FSIM, VSI, HaarPSI, and DISTS) are better correlated with diagnostic quality perceived by radiologists than PSNR and SSIM. This better correlation suggests that visual feature metrics may capture more relevant aspects for diagnostic interpretation than simple pixel-wise comparisons. We computed the metrics for patch evaluation using the PyTorch Image Quality (PIQ) package [49].

#### 2.5.2 SR reconstruction of MRI volumes

We evaluated the quality of SR reconstructed MRI volumes with respect to ground truth HR MRI using the PSNR and SSIM. These two measures were chosen for their straightforward applicability to 3D images. We opted to compute these measures within a brain mask to ensure they accurately reflect the quality of SR in diagnostically relevant areas, avoiding inflation from the uniformity of background air. We also included a comparison with two state-of-the-art methods for structural MRI SR that have publicly available implementations:

- SMORE-v4: the *”Synthetic Multi-Orientation Resolution Enhancement”* [50, 21] method is a single-image SR algorithm devised to increase the through-plane resolution of multi-slice MRI. It is based on the self-training of a CNN model for super-resolution using patches extracted from the HR plane. Then, the self-trained models are applied to LR slices to obtain a HR MRI volume.
- SOUP-GAN: the *”Super-resolution Optimized Using Perceptual-tuned GAN”* [26] is a single-image 3D SR framework to produce thinner slices of MRI. In this work, authors trained a GAN using a perceptual loss calculated from slices in the axial, sagittal, and coronal view.

#### 2.5.3 Impact of SR on automated lesion segmentation

The impact of SR on WM lesion segmentation was evaluated by applying two automated lesion segmentation methods to the SR reconstructed T2-W FLAIR volumes. These two methods were chosen because they accept T2-w FLAIR as the only input and are publicly available:

- **LST-lpa** [51]: the lesion prediction algorithm (lpa) is part of the Lesion Segmentation Toolbox (LST) for SPM. It is a statistical method based on a logistic regression model that includes a lesion belief map and a spatial covariate that takes into account voxel-specific changes in lesion probability.
- **SAMSEG** [52]: the lesion segmentation add-on to SAMSEG routine in Freesurfer, it allows the simultaneous segmentation of white matter lesions and 41 structures by decoupling computational models of anatomy from models of the imaging process.

We applied these two segmentation methods also to the LR and the HR T2-weighted FLAIR images, being the latter a reference benchmark for segmentation performance.

Finally, we included the results of applying the recently proposed WMH-SynthSeg [53] to the simulated LR T2-w FLAIR images. WMH-SynthSeg is an automated method aiming to segment WM hyper-intensities and 36 anatomical brain regions from MRI of any resolution and contrast. Regardless of the input, WMH-SynthSeg produces a HR segmentation volume with 1 mm isotropic voxels, going directly from LR images to HR lesion segmentation.

## 3 Results

### 3.1 SR of MRI patches

The ability of fine-tuned models, namely EDSR and RealESRGAN, to upsample LR MRI patches was evaluated with seven image quality measures: PSNR, SSIM, VIF, FSIM, VSI, HaarPSI, and DISTS. The mean and standard deviation of each image measure along patches extracted from the evaluation set is presented in Table 2. The fine-tuned EDSR is superior to the fine-tuned RealESRGAN in all quality measure for both sequences, T2-W FLAIR and T1-W MRI. Meanwhile, both fine-tuned CNNs outperform bicubic interpolation by at least 1 dB in PSNR and 0.06 in SSIM. Figure 2 shows four examples of super-resolution of LR patches using the fine-tuned models.

**Figure 2:**
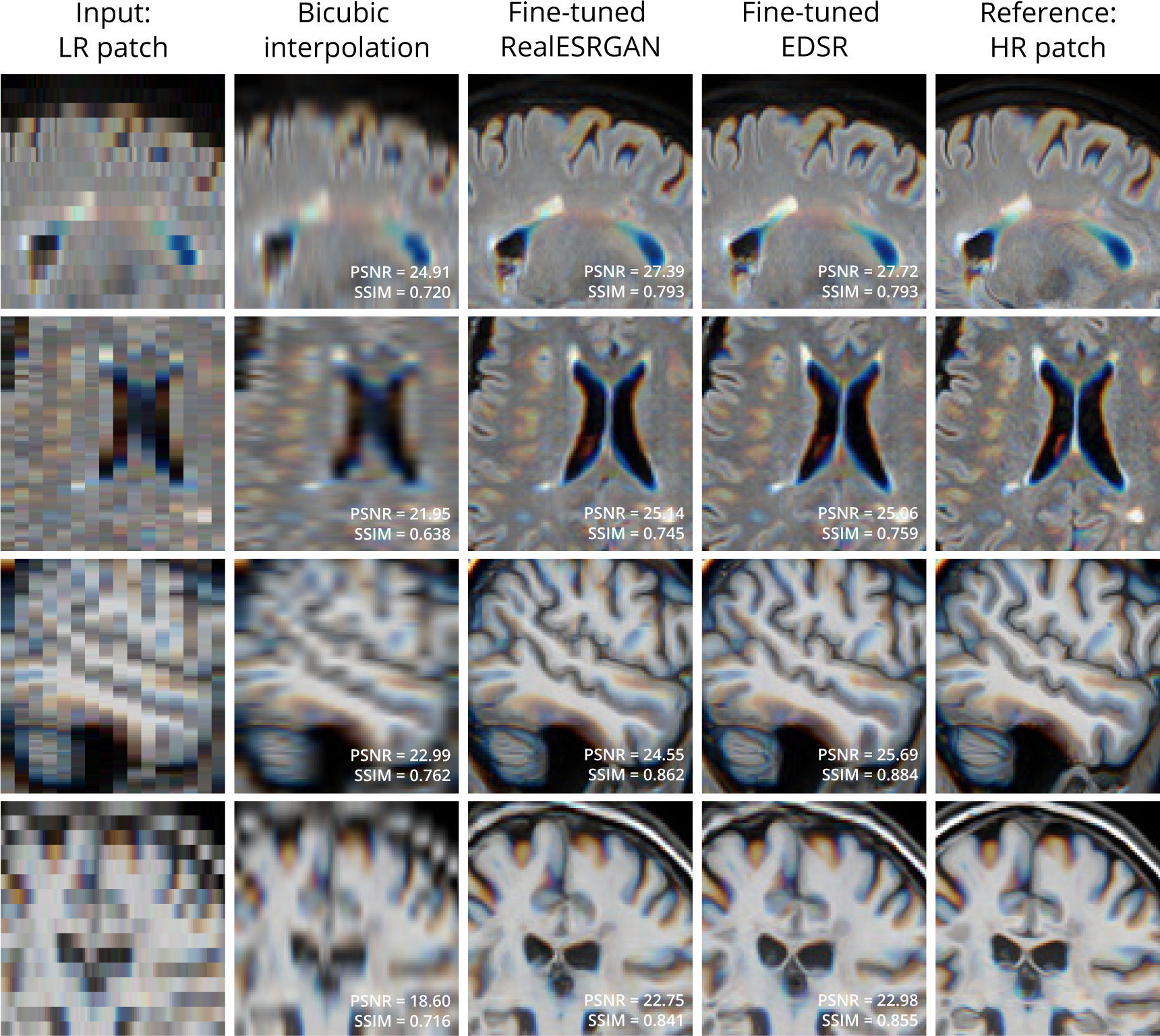
Examples of bicubic interpolation and SR with fine-tuned models applied to LR patches extracted from T2-W FLAIR (first 2 rows) and T1-W MRI (last 2 rows). Sets of three contiguous 2D LR patches are taken as one RGB image.

**Table 2:** Mean and standard deviation of image quality measures (IQM) of fine-tuned CNN models, RealESRGAN and EDSR, per MRI sequence. Results of bicubic interpolation are also included for comparison.

| MRI sequence | IQM | Bicubic interpolation | Fine-tuned RealESRGAN | Fine-tuned EDSR |
| --- | --- | --- | --- | --- |
| T2-W FLAIR | PSNR (dB) | $22.7 \pm 1.2$ | $23.7 \pm 1.2$ | **$24.1 \pm 1.3$** |
| | SSIM | $0.64 \pm 0.06$ | $0.70 \pm 0.05$ | **$0.72 \pm 0.06$** |
| | VIF | $0.28 \pm 0.04$ | $0.33 \pm 0.05$ | **$0.35 \pm 0.05$** |
| | FSIM | $0.82 \pm 0.03$ | $0.87 \pm 0.02$ | **$0.88 \pm 0.02$** |
| | HaarPSI | $0.64 \pm 0.03$ | $0.72 \pm 0.03$ | **$0.74 \pm 0.04$** |
| | VSI | $0.92 \pm 0.01$ | $0.94 \pm 0.01$ | **$0.95 \pm 0.01$** |
| | DISTS | $0.30 \pm 0.02$ | $0.14 \pm 0.02$ | **$0.13 \pm 0.02$** |
| T1-W | PSNR (dB) | $21.0 \pm 2.0$ | $23.0 \pm 1.8$ | **$23.7 \pm 1.9$** |
| | SSIM | $0.71 \pm 0.05$ | $0.79 \pm 0.05$ | **$0.81 \pm 0.05$** |
| | VIF | $0.32 \pm 0.05$ | $0.39 \pm 0.05$ | **$0.41 \pm 0.06$** |
| | FSIM | $0.83 \pm 0.03$ | $0.88 \pm 0.03$ | **$0.89 \pm 0.03$** |
| | HaarPSI | $0.65 \pm 0.05$ | $0.75 \pm 0.04$ | **$0.77 \pm 0.04$** |
| | VSI | $0.93 \pm 0.02$ | $0.95 \pm 0.01$ | **$0.96 \pm 0.01$** |
| | DISTS | $0.28 \pm 0.03$ | $0.13 \pm 0.02$ | **$0.12 \pm 0.02$** |

Compared to bicubic interpolation, notable improvements are evident in tissue boundaries, sulci shape, and lesion surroundings. It should be noted that MRI patches for evaluation were extracted following the same approach used for training: three contiguous 2D LR patches taken as one RGB image. An extended comparison including examples with the pre-trained models is shown in Supplementary material.

### 3.2 SR reconstruction of MRI volumes

The quality of SR reconstructed MRIs was evaluated with the 3D versions of PSNR and SSIM, calculated within a brain mask. Table 3 presents the mean and standard deviation of these metrics for our SR framework, PRETTIER, compared to SMORE and SOUP-GAN. Our approach using the fine-tuned EDSR (PRETTIER-EDSR), consistently outperforms all other methods across all MRI sequences and evaluation sets (see Supplementary material for distributions per dataset). PRETTIER with the fine-tuned RealESRGAN yields higher PSNR and SSIM than SMORE when reconstructing T2-W FLAIR images but lower when reconstructing T1-W MRI. Notably, both PRETTIER and SMORE substantially outperform SOUP-GAN.

**Table 3:**
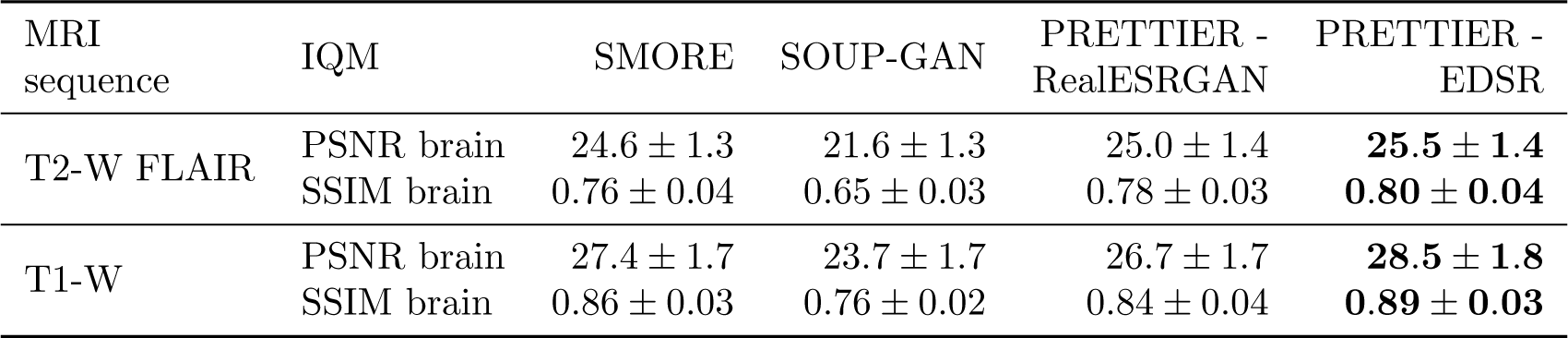
Mean and standard deviation of PSNR and SSIM for MRI SR reconstruction methods, calculated within a brain mask.

| MRI sequence | IQM | SMORE | SOUP-GAN | PRETTIER - RealESRGAN | PRETTIER - EDSR |
| --- | --- | --- | --- | --- | --- |
| T2-W FLAIR | PSNR brain | $24.6 \pm 1.3$ | $21.6 \pm 1.3$ | $25.0 \pm 1.4$ | **$25.5 \pm 1.4$** |
| | SSIM brain | $0.76 \pm 0.04$ | $0.65 \pm 0.03$ | $0.78 \pm 0.03$ | **$0.80 \pm 0.04$** |
| T1-W | PSNR brain | $27.4 \pm 1.7$ | $23.7 \pm 1.7$ | $26.7 \pm 1.7$ | **$28.5 \pm 1.8$** |
| | SSIM brain | $0.86 \pm 0.03$ | $0.76 \pm 0.02$ | $0.84 \pm 0.04$ | **$0.89 \pm 0.03$** |

Qualitative comparisons for T2-W FLAIR and T1-W are shown in Figure 3 and Figure 4, respectively. While quantitative results indicate a relatively modest increase of PSNR and SSIM over SMORE, qualitative results show noticeable improvements such as better-defined lesion contours in T2-W FLAIR (Figure 3) and more anatomically coherent tissue boundaries in T1-W MRI (Figure 4). Meanwhile, SOUP-GAN appears to introduce artifacts and textures that are not present in the ground truth HR image, which might explain its lower performance metrics compared to the other methods.

**Figure 3:**
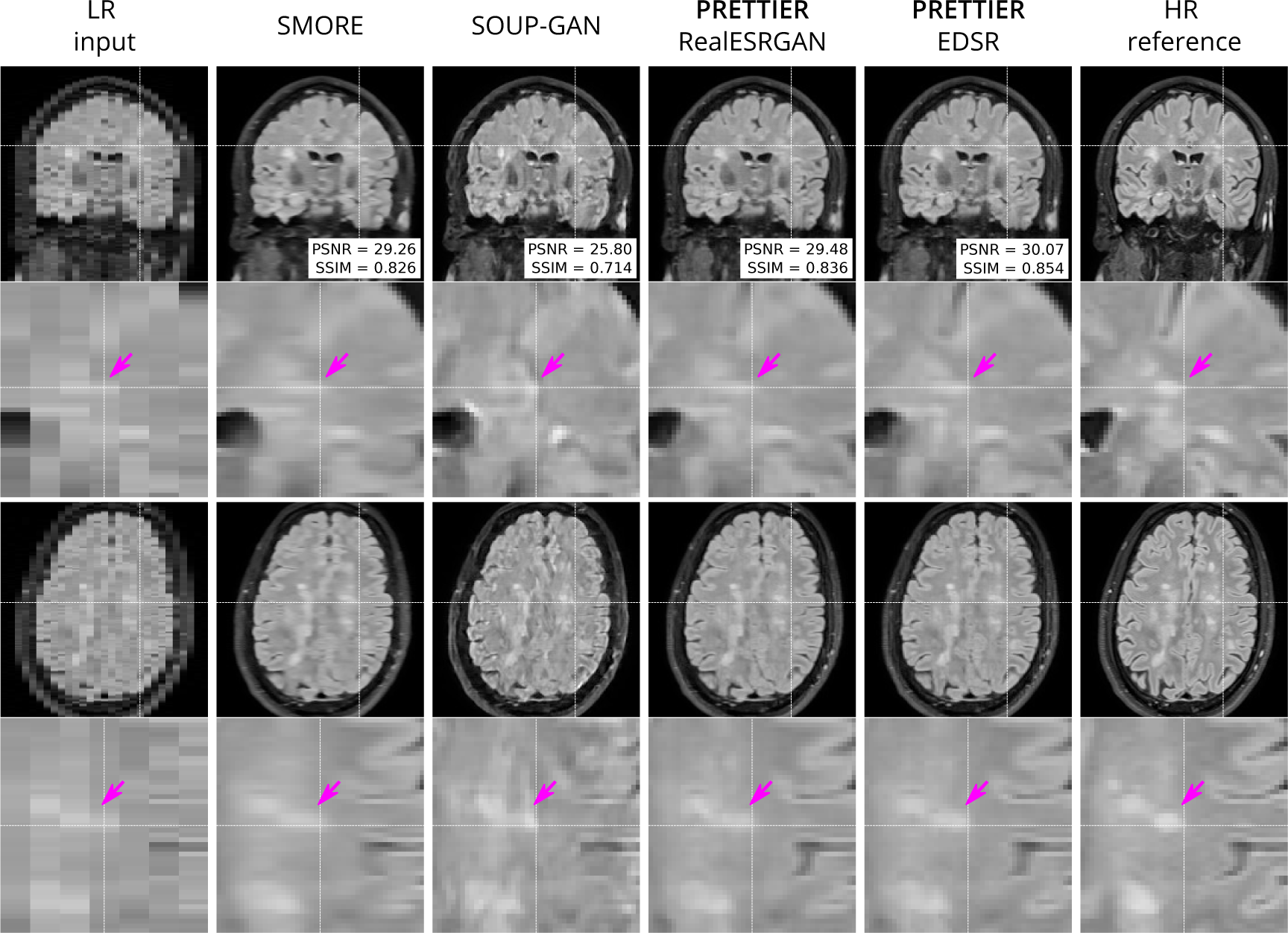
Qualitative result for simulated LR T2-W FLAIR with sagittal slice orientation. Coronal (top row) and axial (bottom row) views of LR input, volumes reconstructed using SMORE [50, 21], SOUP-GAN [26], and our SR framework (PRETTIER) with the fine-tuned RealESRGAN and EDSR, and the HR reference volume. PSNR and SSIM values are calculated within a brain mask.

**Figure 4:**
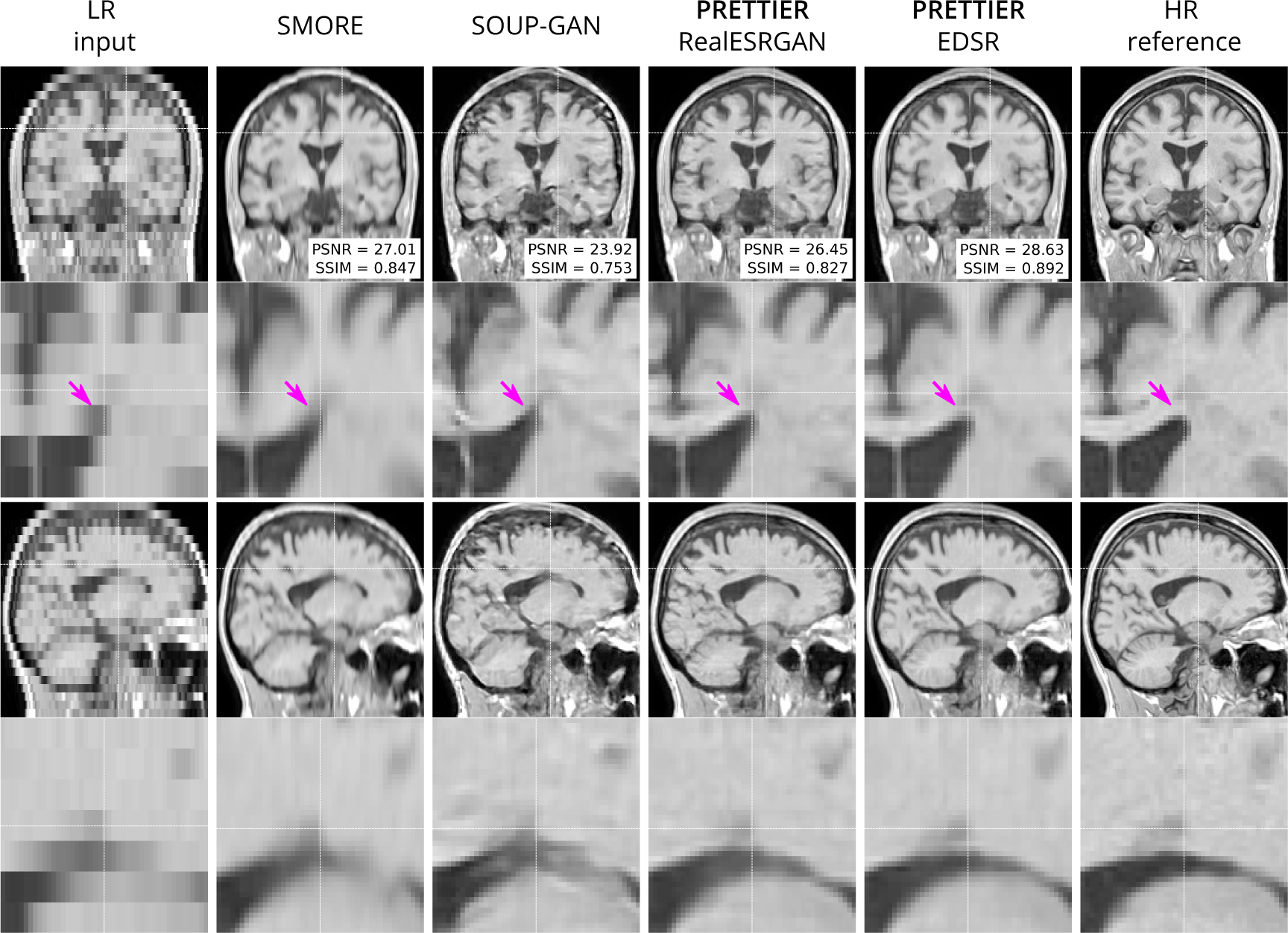
Qualitative result for simulated low-resolution LR T1-W MRI with axial slice orientation. Coronal (top row) and sagittal (bottom row) views of LR input, volumes reconstructed using SMORE [50, 21], SOUP-GAN [26], and out SR framework (PRETTIER) with the fine-tuned RealESRGAN and EDSR, and the HR reference volume. PSNR and SSIM values are calculated within a brain mask.

### 3.3 Effect of SR on automated lesion segmentation

The performance of automated lesion segmentation was quantitatively evaluated in terms of the Dice score, sensitivity, precision, and error of lesion volume estimation. In Table 4, we present the mean and standard deviation of the first three segmentation performance measures for LST-lpa and SAMSEG when applied to LR, SR reconstructed, and HR T2-W FLAIR images, and for WMH-SynthSeg applied only to LR images. Due to non-gaussianity and presence of outliers, we present the distributions of lesion volume estimation errors in Figure 5, a similar plot for Dice score, sensitivity, and precision is presented in the Supplementary material. These results confirm that applying PRETTIER improves lesion segmentation over LR images and brings it closer to segmentation performance in ground truth HR images. Furthermore, in the scenario of segmenting WM lesions on LR multi-slice T2-W FLAIR of PwMS, a better Dice score is achieved by applying first PRETTIER-EDSR and then segmenting lesions with LST-lpa than by applying WMH-SynthSeg directly on the LR image.

**Figure 5:**
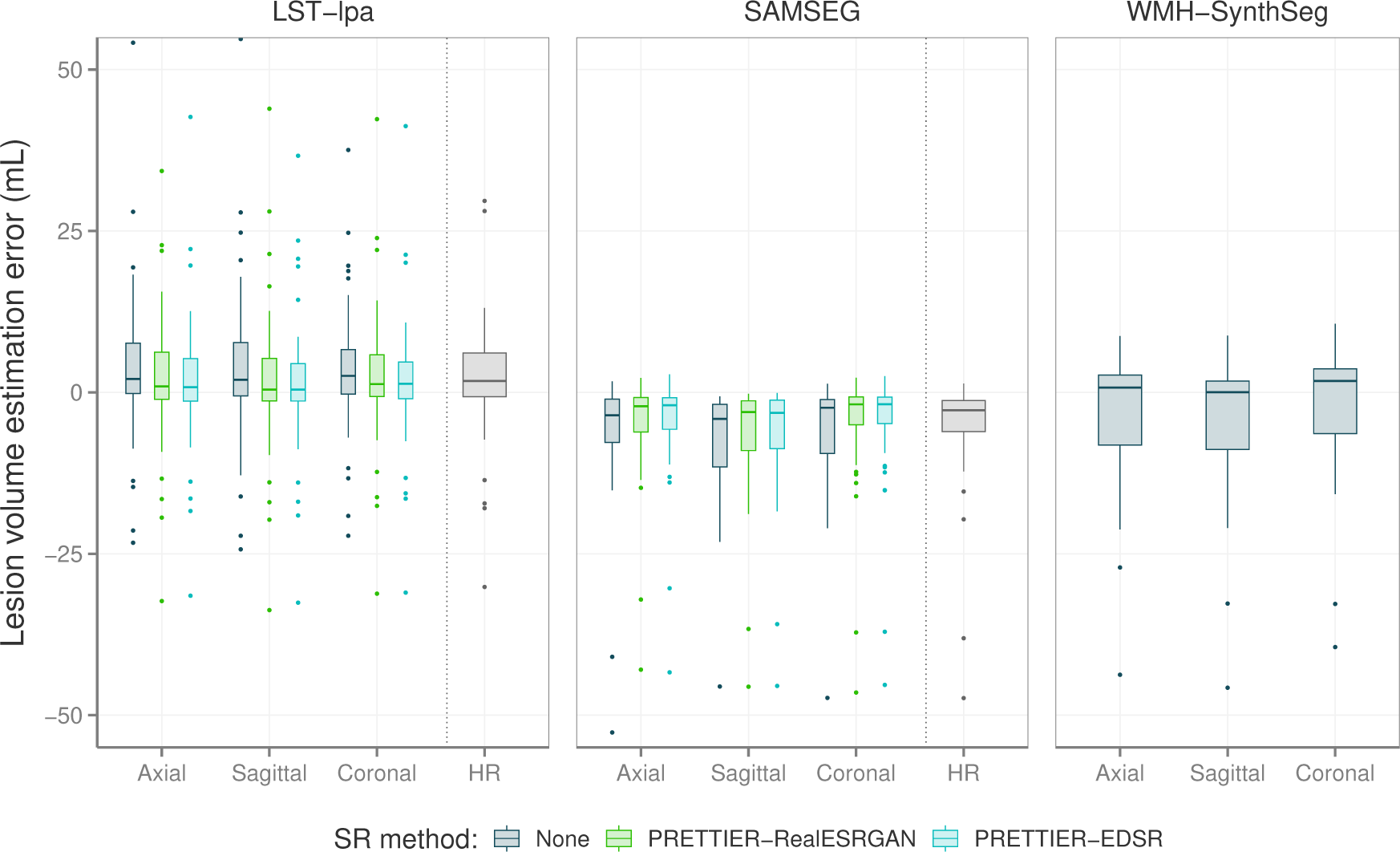
Distribution of error in lesion volume estimation from automated segmentation with LST-lpa [51], SAMSEG [52], and WMH-SynthSeg [53].

**Table 4:** Mean and standard deviation of Dice score, sensitivity, and precision calculated for automated lesion segmentation on T2-W FLAIR. LST-lpa [51] and SAMSEG [52] were applied to simulated LR images, the outputs of our SR framework, and to ground truth HR images. WMH-SynthSeg [53] was only applied to LR images as its purpose is to directly produce HR segmentations.

| Method | Input | Dice score | Sensitivity | Precision |
| --- | --- | --- | --- | --- |
| LST-lpa | LR | $0.47 \pm 0.15$ | $0.59 \pm 0.17$ | $0.47 \pm 0.22$ |
| | PRETTIER-RealESRGAN | $0.52 \pm 0.15$ | $0.60 \pm 0.19$ | $0.55 \pm 0.22$ |
| | PRETTIER-EDSR | $0.53 \pm 0.15$ | $0.60 \pm 0.18$ | $0.55 \pm 0.22$ |
| | HR | $0.58 \pm 0.16$ | $0.69 \pm 0.16$ | $0.57 \pm 0.23$ |
| SAMSEG | LR | $0.41 \pm 0.17$ | $0.32 \pm 0.16$ | $0.67 \pm 0.16$ |
| | PRETTIER-RealESRGAN | $0.46 \pm 0.21$ | $0.38 \pm 0.21$ | $0.71 \pm 0.16$ |
| | PRETTIER-EDSR | $0.47 \pm 0.21$ | $0.40 \pm 0.21$ | $0.71 \pm 0.17$ |
| | HR | $0.53 \pm 0.20$ | $0.43 \pm 0.19$ | $0.79 \pm 0.14$ |
| WMH-SynthSeg | LR | $0.35 \pm 0.14$ | $0.37 \pm 0.14$ | $0.40 \pm 0.23$ |

Figure 6 shows an example of WM lesion segmentation on T2-W FLAIR, applied to LR inputs, our SR reconstructions, and HR images. This example illustrates how applying SR enhances the automated lesion segmentation when the input is a LR image (red mask in the figure). The automated segmentation on the HR image (green mask) serves as an upper bound, indicating the best segmentation performance attainable for each method, which might still be far from the ground truth segmentation (blue mask).

**Figure 6:**
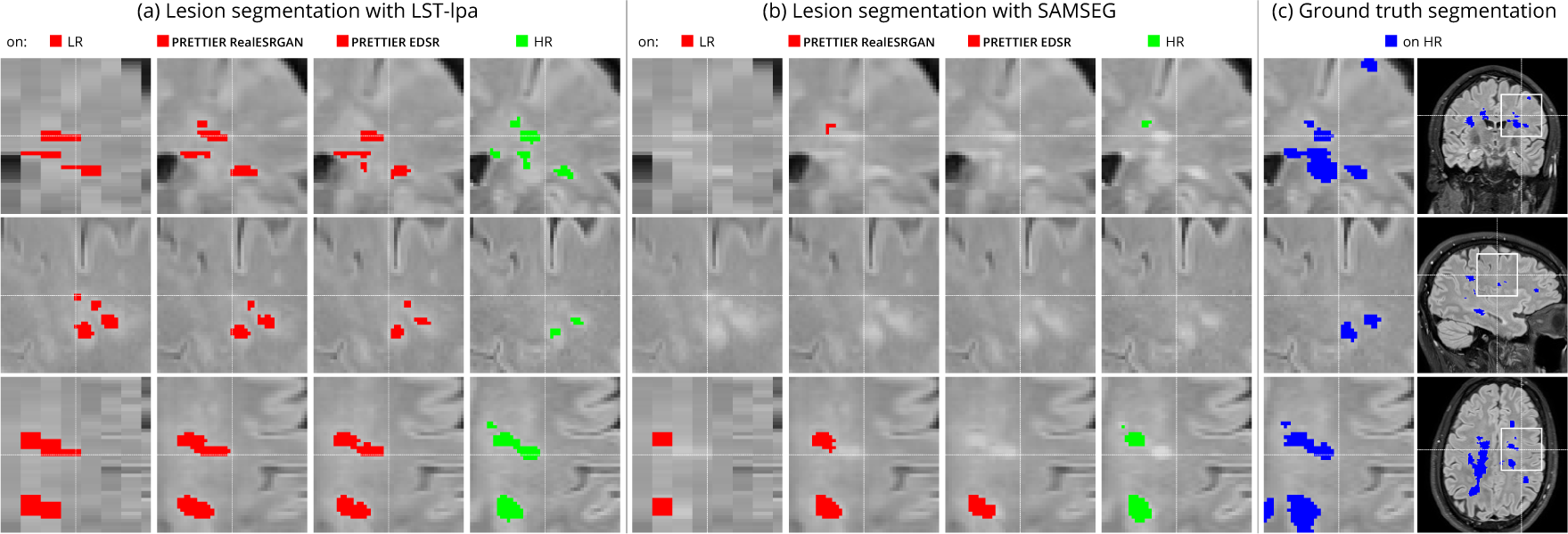
Example of automated white matter lesion segmentation (a) LST-lpa and (b) SAMSEG, compared against (c) the ground truth manual segmentation. Our SR framework improves the segmentation over LR T2-W FLAIR (red), approaching the performance achieved with the highresolution image (green).

## 4 Discussion

In this work, we presented PRETTIER, a framework to enhance the through-plane resolution of multi-slice structural MRIs containing MS lesions. Evaluation results with independent datasets demonstrate successful SR reconstruction which leads to improved performance of automated lesion segmentation.

There are four key aspects that contribute to the successful domain adaptation of 2D CNN models in our approach. First, the content loss function guiding the fine-tuning process promotes not only pixel-wise reconstruction accuracy but also the recovery of high-level features by including the perceptual loss. This loss formulation leads to outputs with high perceptual quality, as shown in the evaluation with metrics beyond PSNR and SSIM (See Table 2), as well as in the qualitative results exhibiting well defined tissue and lesion boundaries (See Figure 2 and Figure 1 in Supplementary material). Second, we use patches extracted from two different MRI modalities (T2-W FLAIR and T1-W MRI) with three different slice orientations (axial, sagittal, and coronal). This variability in our training dataset exposes the models to a wide range of anatomical and contrast variations, potentially enhancing their generalizability, as suggested by the results in our similarly diverse evaluation dataset. Third, instead of of using simple downsampling or k-space truncation as done in some existing literature [17, 18, 19], we obtain pairs of LR-HR images by applying a physics-informed model of multi-slice MRI acquisition to HR images which takes into account the slice selection profile. Fourth, we incorporate information from adjacent slices as color channels in the inputs and outputs of 2D CNN models, allowing them to leverage the 3D information in MRI while benefiting from architectural advances in natural image SR.

Two considerations led us to employ 2D instead of 3D CNN architectures. First, 2D models have lower computational costs during training and inference compared to their 3D counterparts. Second, adopting a 2D approach offers greater flexibility in leveraging advances from the vast literature on natural image SR. Many of the cutting-edge architectures and strategies for SR have been primarily developed and optimized for 2D images. By operating with 2D models, we could adapt and fine-tune any of these models to our MRI SR framework. Specifically, in this work we used two SR models: the EDSR [14] and the RealESRGAN [16]. Patch-based evaluation of finetuned models demonstrates that EDSR consistently outperforms RealESRGAN across datasets, MRI contrasts and evaluation metrics (See Table 2). It is worth noting the architectural differences between these models. EDSR has 32 residual blocks with 256 features in each convolutional layer, amounting to over 40 million trainable parameters. Meanwhile, the generator in RealESRGAN has 23 residual-in-residual dense blocks with 64 initial features per residual dense block, resulting in approximately 16.7 million parameters, a lighter model size than EDSR.

We quantitatively evaluate our framework for SR reconstruction of MRI volumes using PSNR and SSIM metrics, comparing against two state-of-the-art methods for MRI SR: SMORE [50, 21] and SOUP-GAN [26]. The key feature of SMORE is its self-supervised training without relying on external data. However, this comes at a significant computational cost as a CNN model must be trained for each new input image. In contrast, our approach leverages trained models that have been fine-tuned with data we gathered from external datasets, allowing faster and less resource demanding application. Evaluation on the independent dataset shows our fine-tuned EDSR model outperforms SMORE across metrics for both T2-W FLAIR and T1-W MRI (See Table 3 and Figure S2 in Supplementary material). While those quantitative results also show our approach with the fine-tuned RealESRGAN slightly underperforms SMORE on T1-W, the qualitative example (Figure 4) reveals sharper tissue boundaries more alike the HR ground truth. Our SR framework shares some similarities with SOUP-GAN [26], as both approaches rely on models trained with a perceptual loss. Specifically, SOUP-GAN employs a scale-attention architecture trained via an adversarial approach. However, our evaluation results demonstrate that SOUP-GAN underperforms quantitatively and qualitatively compared to both SMORE and our framework using the fine-tuned EDSR and RealESRGAN models. The qualitative examples in Figure 3 and Figure 4 suggest that SOUP-GAN suffers from artificial textures, artifacts that are likely introduced when promoting only the perceptual quality of images (via perceptual loss and adversarial training) without accounting for reconstruction accuracy.

White matter lesion segmentation is a highly relevant task when processing brain MRI data of PwMS. Assessing the impact of our SR framework on this task is crucial for validating its practical use. For this evaluation, we applied two different automated methods for lesion segmentation, LST-lpa [51] and SAMSEG [52], on T2-W FLAIR images. We compared the segmentation performance when using the LR images versus the SR reconstructed images, and also include the segmentation performance on HR images as reference. The results demonstrate that, compared to segmentation on LR images, our SR reconstruction approach improves the Dice score, sensitivity, precision, and lesion volume estimation, bringing them closer to what is achievable with HR images (Table 4 and Figure 5). Consistently, we observe that LST-lpa exhibits higher sensitivity but lower precision than SAMSEG when applied to LR and HR T2-W FLAIR images. Notably, our SR approach improves the precision of LST-lpa without compromising sensitivity, suggesting it effectively refines lesion boundaries, as illustrated in Figure 6. Conversely, SR enhances the low sensitivity of SAMSEG while also improving its precision. Furthermore, we also include a comparison against the recently proposed WMH-SynthSeg [53], a method aiming to produce a HR segmentations of WM hyperintensities (and 36 brain regions) from scans of any resolution and contrast. Our evaluation shows that, given a LR T2-W FLAIR image (acquired in a 1.5T or 3T scanner), applying our SR approach followed by LST-lpa or SAMSEG yields superior lesion segmentation compared to directly applying WMH-SynthSeg on the LR images.

The work presented herein has some limitations. First, LR MRIs used for model fine-tuning and evaluation are simulated using only one slice profile: slice thickness of 5 mm and slice spacing of 6 mm (*i.e.*, 1 mm of slice gap). While our evaluation results demonstrate the capabilities of our SR framework in this challenging and common clinical scenario, and preliminary results show good performance on images with different slice profiles (See Supplementary Figure), future work should evaluate its performance across a broader range of acquisition settings. Second, the computational requirements of using deep CNN models can pose barriers to their implementation, especially in resource-limited settings. To address this, future research will explore the capabilities of more efficient SR models [54, 55]. Lastly, we evaluated the impact of our SR framework on only one downstream task, the automated segmentation of WM lesions. Expanding this evaluation to other downstream tasks in MS neuroimaging analyses, such as regional volumetry and radiomic feature extraction, would provide a more comprehensive assessment of SR potential benefits and limitations in MS research.

In conclusion, we have presented PRETTIER, a single-image SR framework for multi-slice structural MRI of PwMS that leverages existing CNN architectures for image SR. Our framework demonstrates superior image quality results than existing methods for MRI SR, and improves the automated lesion segmentation on LR T2-W FLAIR. By effectively addressing the limitations of routinely acquired multi-slice MRI with low through-plane resolution, our approach facilitates the use of retrospective MRI datasets already acquired in the clinics to conduct 3D analyses and investigate image-based biomarkers of MS outcomes.

## Conflict of Interest Statement

The authors declare that the research was conducted in the absence of any commercial or financial relationships that could be construed as a potential conflict of interest.

## Funding

This research received funding from the Flemish Government under the “Onderzoeksprogramma Artificïele Intelligentie (AI) Vlaanderen” program.

## Acknowledgments

We gratefully acknowledge the collaboration of the ”Centro de Esclerosis Múltiple del Hospital Universitario Nacional de Colombia” (https://www.hun.edu.co/CEMHUN) in the construction of the HUN dataset.

The MSSEG1 and MSSEG2 datasets were made available by The Observatoire Fraņcais de la Scĺerose en Plaques (OFSEP), who is supported by a grant provided by the French State and handled by the “Agence Nationale de la Recherche,” within the framework of the “Investments for the Future” program, under the reference ANR-10-COHO-002, by the Eugène Devic EDMUS Foundation against multiple sclerosis and by the ARSEP Foundation.

## Data Availability Statement

The code to apply the presented super-resolution framework for structural MRI is available at https://github.com/diagiraldo/PRETTIER.

Data used in this work included four publicly available datasets: ISBI2015 [29], Lesjak-3D [30], MSSEG1 [31], and MSSEG2 [32]. Two clinical datasets, MSPELT and HUN, are not openly available due to privacy concerns.

## Footnotes

1 https://github.com/sanghyun-son/EDSR-PyTorch/

2 https://github.com/xinntao/Real-ESRGAN/

## Notes

### Competing Interest Statement

The authors have declared no competing interest.

### Funding Statement

This research received funding from the Flemish Government under the 'Onderzoeksprogramma Artificiäle Intelligentie (AI) Vlaanderen program'.

### Author Declarations

Ethical commission of the University of Hasselt gave ethical approval for this work (CME2019/046). Ethics committee of Hospital Universitario Nacional de Colombia gave ethical approval for this work (CEI-HUN-2019-12).

