## Supplementary Material for "Perceptual super-resolution in multiple sclerosis MRI"

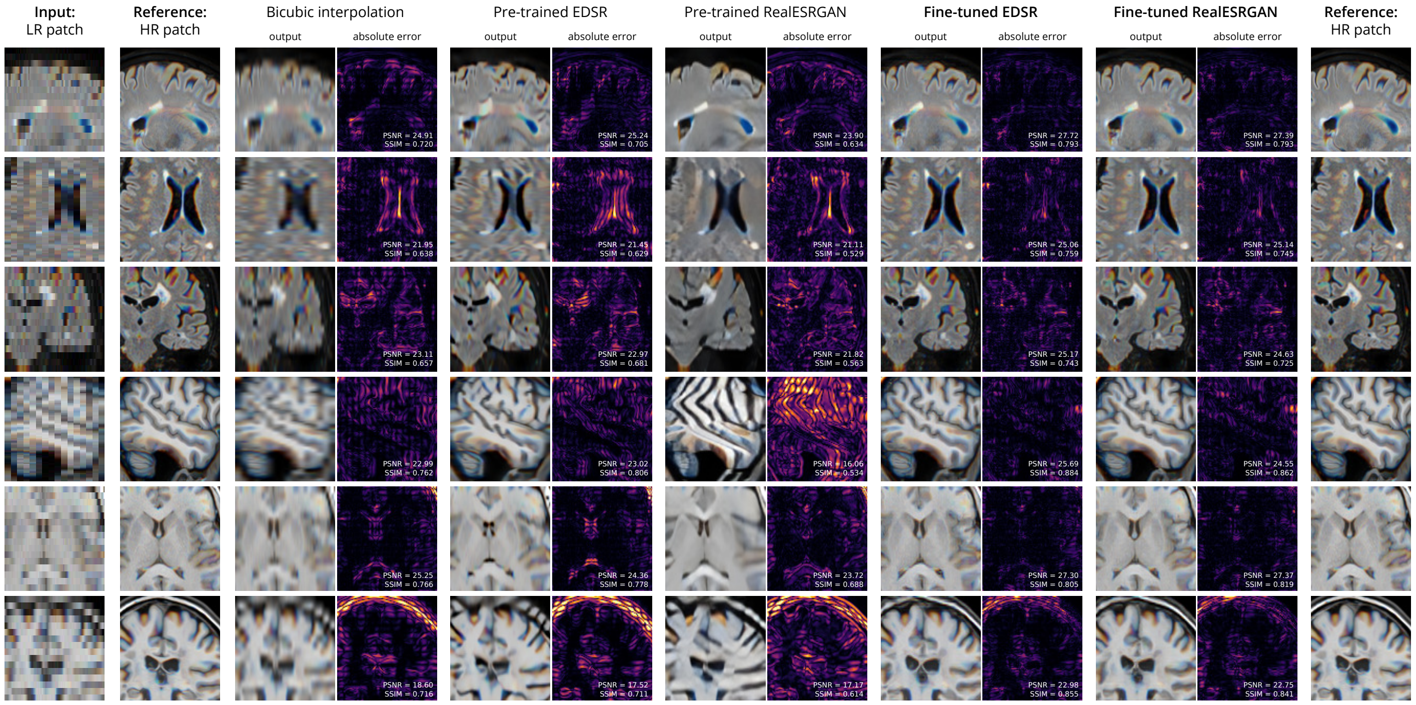


Figure 1: Examples of paired LR-HR patches, bicubic interpolation applied to LR patches, and patch super-resolution with pre-trained and fine-tuned CNN models.

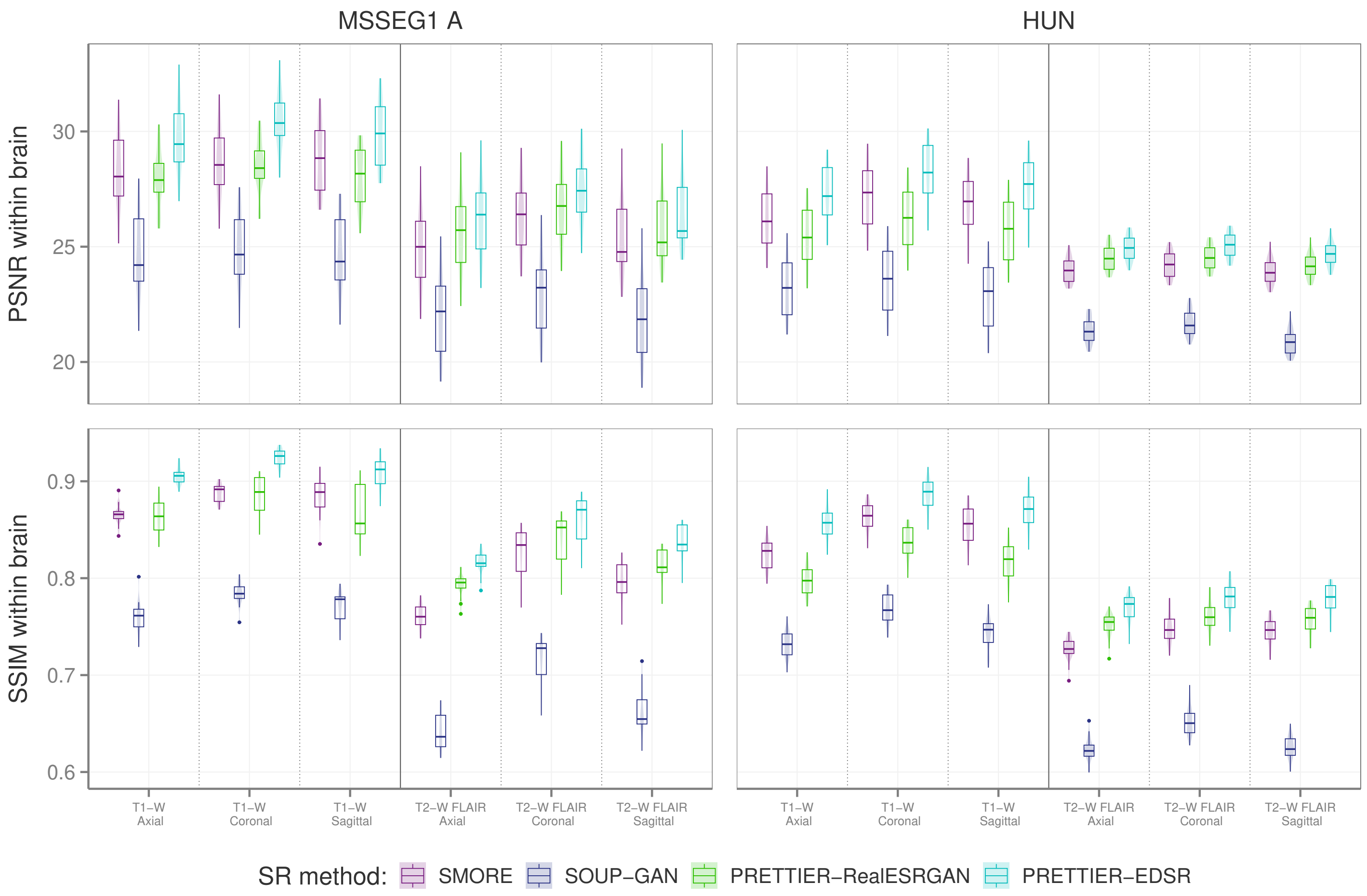


Figure 2: PSNR and SSIM distributions for reconstruction of MRI volumes in the evaluation set.

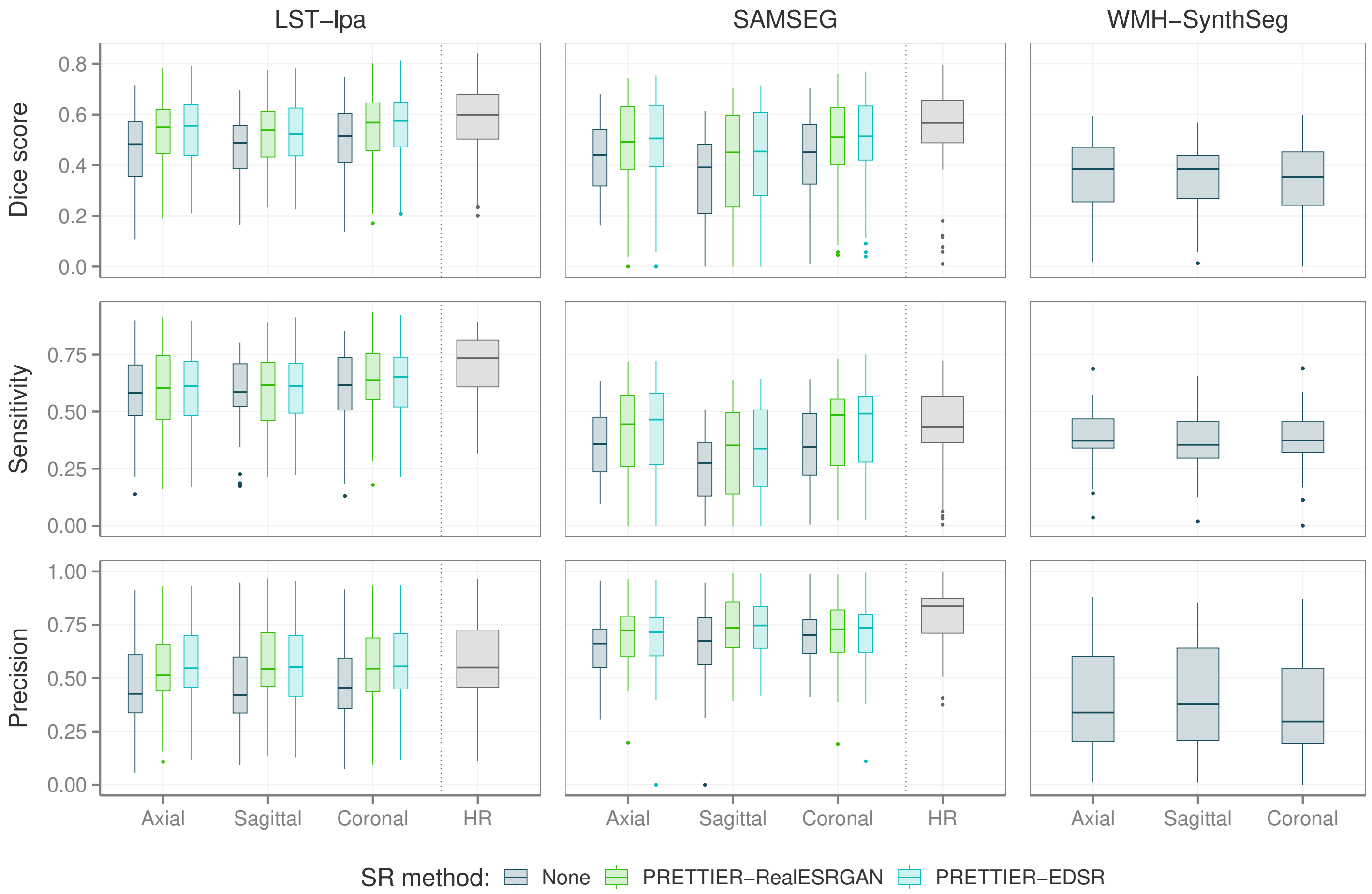


Figure 3: Distributions of Dice score, sensitivity, and precision for automated lesion segmentation on T2-W FLAIR images with LST-lpa, SAMSEG and WMH-SynthSeg.

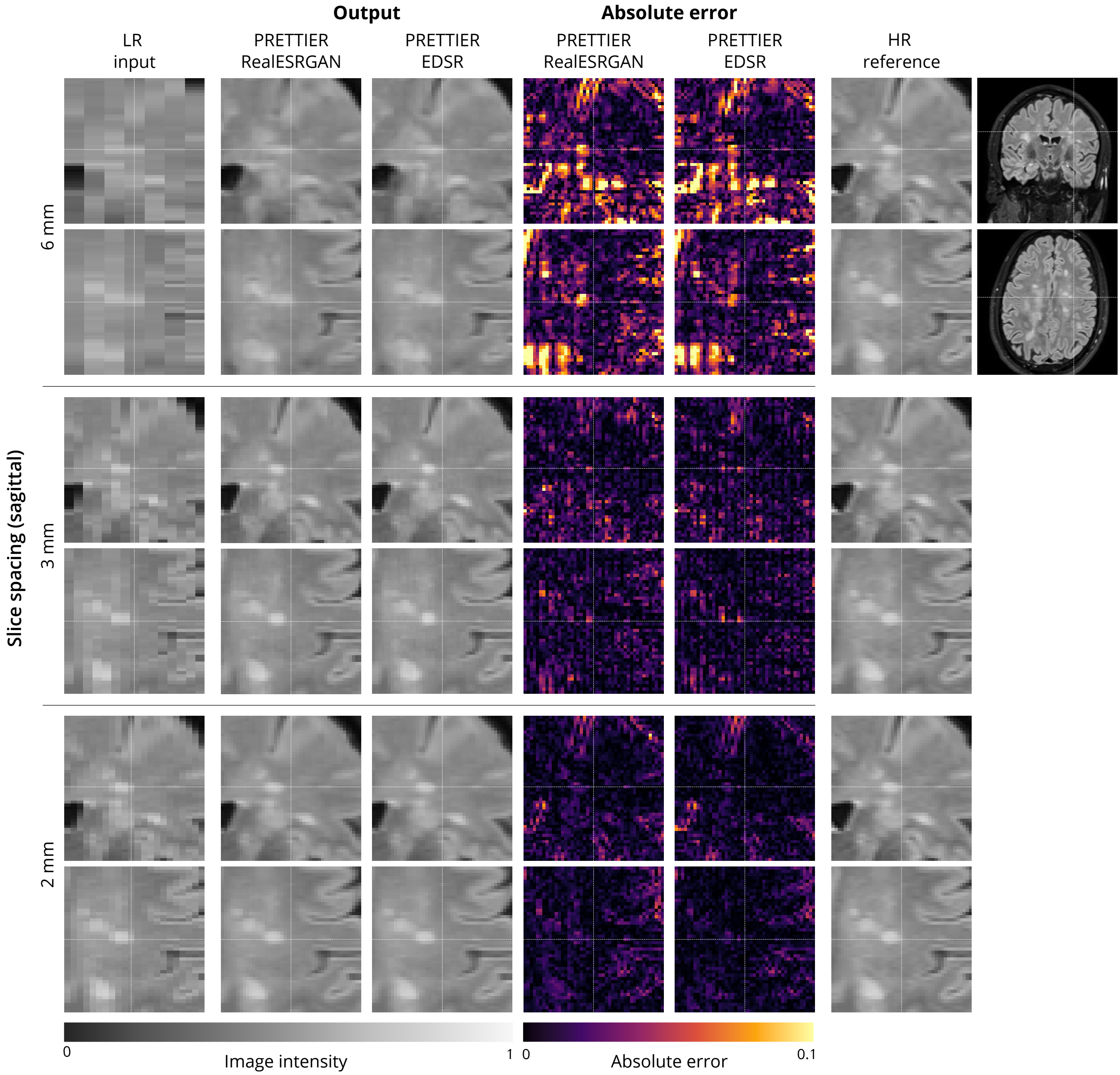


Figure 4: Qualitative results of PRETTIER SR reconstructions from LR T2-W FLAIR, simulated with sagittal slice orientation and three different slice profiles: 6 mm of slice spacing with 1 mm of slice gap (top panel), 3 mm of slice spacing without slice gap (middle panel), and 2 mm of slice spacing without slice gap (bottom panel).

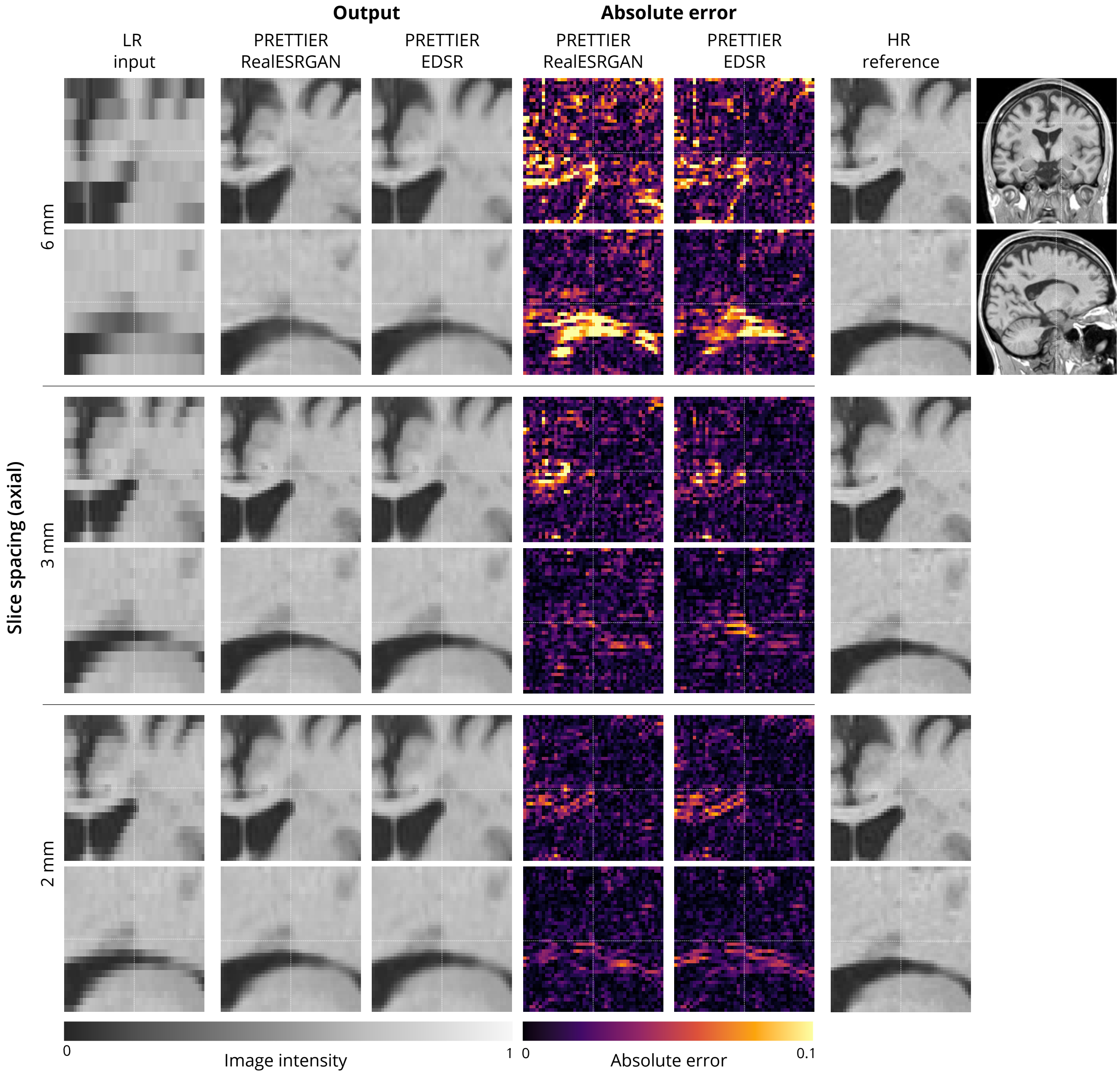


Figure 5: Qualitative results of PRETTIER SR reconstructions from LR T1-W MRI, simulated with axial slice orientation and three different slice profiles: 6 mm of slice spacing with 1 mm of slice gap (top panel), 3 mm of slice spacing without slice gap (middle panel), and 2 mm of slice spacing without slice gap (bottom panel).
